# Characteristics and Outcomes of Multisystem Inflammatory Syndrome in Children: A Multicenter, Retrospective, Observational Cohort Study in Mexico

**DOI:** 10.1101/2023.02.16.23285979

**Authors:** Nadia González-García, Marco Antonio Yamazaki-Nakashimada, Horacio Márquez-González, Guadalupe Miranda-Novales, Gonzalo Antonio Neme Díaz, Sandhi Anel Prado Duran, Antonio Luévanos Velázquez, Maria F. Castilla-Peon, Miguel Alejandro Sánchez Duran, Martha Patricia Márquez Aguirre, Miguel Angel Villasis-Keever, Ranferi Aragón Nogales, Carlos Núñez Enríquez, Maria Elena Martinez Bustamante, Carlos Aguilar Argüello, Jesús Ramírez de los Santos, Alejandra Pérez Barrera, Lourdes Anais Palacios Cantú, Jesús Membrila Mondragón, Paloma Vizcarra Alvarado, Rodolfo Norberto Jiménez Juárez, Víctor Olivar López, Adrián López Chávez

## Abstract

Multisystem inflammatory syndrome in children temporally associated with coronavirus disease 2019 (MIS-C), a novel hyperinflammatory condition secondary to severe acute respiratory syndrome coronavirus 2 (SARS-CoV-2) infection, is associated with severe outcomes such as coronary artery aneurysm and death. This multicenter, retrospective, observational cohort study including eight centers in Mexico, aimed to describe the clinical characteristics and outcomes of patients with MIS-C. Patient data were evaluated using latent class analysis to categorize patients into three phenotypes: toxic shock syndrome-like (TSSL)-MIS-C, Kawasaki disease-like (KDL)-MIS-C, and nonspecific MIS-C (NS-MIS-C). Risk factors for adverse outcomes were estimated using multilevel mixed-effects logistic regression. The study included 239 patients with MIS-C, including 61 (26%), 70 (29%), and 108 (45%) patients in the TSSL-MIS-C, KDL-MIS-C, and NS-MIS-C groups, respectively. Fifty-four percent of the patients were admitted to the intensive care unit, and 42%, 78%, and 41% received intravenous immunoglobulin, systemic glucocorticoids, and anticoagulants, respectively. Coronary artery dilatation and aneurysm were found in 5.7% and 13.2% of the patients, respectively. The rate of mortality due to SARS-CoV-2-related factors was 4.6%. Delay of ≥10 days in hospital admission was associated with coronary artery aneurysm or dilatation (odds ratio [OR] 1.6, 95% confidence interval [CI] 1.2–2.0). Age ≥ 10 years (OR 5.6, 95% CI 1.4–2.04), severe underlying condition (OR 9.3, 95% CI 2.8–31.0), platelet count < 150,000/mm^3^ (OR 4.2, 95% CI 1.2–14.7), international normalized ratio > 1.2 at admission (OR 3.8, 95% CI 1.05–13.9), and serum ferritin concentration > 1500 mg/dL (OR 52, 95% CI 5.9–463) were risk factors for death.

## 1 Introduction

Multisystem Inflammatory Syndrome in Children (MIS-C), temporally associated with coronavirus disease 2019 (COVID-19) also known as pediatric multisystem inflammatory syndrome, is a novel hyperinflammatory condition secondary to severe acute respiratory syndrome coronavirus 2 (SARS-CoV-2) infection and shares features with Kawasaki disease, toxic shock syndrome, and acute COVID-19. MIS-C was first described in April 2020 in the United Kingdom, followed by case reports in other countries. (1,2)

The clinical case definition of MIS-C varies slightly among different health agencies. (2–4) Clinical features include fever, gastrointestinal symptoms, conjunctival injection, rash, and elevated inflammatory markers. (5)Potential complications include shock, multi-organ failure, and myocardial and coronary artery involvement similar to those observed in Kawasaki disease. (6–8) Many patients with MIS-C have evidence of SARS-CoV-2 infection within several weeks before disease onset. (9–11) The estimated incidence of MIS-C is 3–5 per 10,000 among individuals aged <21 years who are infected with SARS-CoV2. Studies in ethnically diverse countries reported a higher incidence of MIS-C in Hispanic and Afro-descendant individuals, emphasizing the need to characterize the clinical spectrum of MIS-C in specific populations. (12)

Several reports have suggested the existence of different MIS-C phenotypes, including the Kawasaki disease-like (KDL)-MIS-C, the toxic shock syndrome-like (TSSL)-MIS-C, as well as the nonspecific MIS-C with predominantly acute respiratory involvement (NS-MIS-C). Differentiating MIS-C from acute COVID-19 and other hyperinflammatory conditions can be challenging for healthcare providers. The treatment approach for MIS-C includes intravenous immunoglobulin (IVIG), systemic glucocorticoids, and anti-inflammatory biological agents; the efficacy of these treatments relies on the timely diagnosis of MIS-C.

Despite its low prevalence, MIS-C is a serious life-threatening condition. Therefore, this study aimed to examine the clinical characteristics and outcomes of KDL-MIS-C, TSSL-MIS-C, and NS-MIS-C among patients with MIS-C in eight tertiary-care pediatric centers in Mexico. We also performed exploratory analyses to identify factors associated with coronary artery abnormalities and death.

## 2. Materials and Methods

### 2.1 Study design

This was a multicenter, retrospective, observational cohort study including patients diagnosed with MIS-C in one of the eight participating pediatric tertiary care centers in Mexico. Four centers were located in Mexico City: Federico Gómez Mexico Children’s Hospital, National Institute of Pediatrics, Siglo XXI Medical Center’s Pediatrics Hospital, and. Guadalajara Civil Hospital was located in Guadalajara, Jalisco; Villa Hermosa Children’s Hospital was located in Villa Hermosa, Tabasco, and one investigator collected data on cases from the state of Tamaulipas.

The study was approved by Federico Gómez Mexico Children’s Hospital Institutional Review Board (approval no. HIM-2022-015) and the local ethics review board of the remaining participating centers.

Probable MIS-C cases were identified by the review of medical records of patients admitted to the emergency room, intensive care unit (ICU), or the general hospital ward for COVID-19. Patient files were evaluated to determine if they met the MIS-C case definition of the Centers for Disease Control and Prevention (Chart 1). (2)

Local investigators at participating centers collected data from the medical records using REDCap (Research Electronic Data Capture) software hosted at Federico Gómez Mexico Children’s Hospital. Personal identifiers were kept confidential at participating centers and were not registered in the general database. Data were retrieved from REDCap, inconsistencies and missing data were corrected through direct communication with local investigators, units of measurement were unified, and variables were converted as necessary.

The worst value measured within the first three days of hospitalization was obtained for laboratory parameters with more than one measurement. Life support measures (respiratory support and vasopressor utilization) and the use of IVIG, systemic glucocorticoids, and anticoagulant therapy were documented as primary therapeutic interventions. Outcomes of interest were admission, length of ICU stay and hospitalization, need for invasive mechanical ventilation, vasopressor support, myocardial depression (left ventricular ejection fraction < 55%), coronary aneurysm (coronary artery diameter z-score ≥ 2.5) and coronary artery dilatation (diameter z-score >2–<2.5) on the echocardiographic evaluation performed during hospitalization, and in-hospital mortality.

In the present study, severe respiratory involvement was defined as the presence of infiltrates on chest X-ray or computed tomography plus the need for oxygen administration with a non-rebreathing mask or a device with higher oxygen concentration. Additionally, “gastrointestinal symptoms” was defined as the presence of at least one of the following: abdominal pain, diarrhea, and emesis. Severe underlying conditions included cancer and other immunosuppressive states, neuromuscular disability, chronic respiratory diseases except for asthma, congenital cardiac disorders, and chronic kidney failure.

### 2.2 Statistical analysis

Descriptive analyses were conducted using STATA v.14.0 (StataCorp, Houston, TX, USA). Categorical variables were reported as frequencies, and continuous variables were reported as medians with interquartile ranges.

In the present study, we utilized LCA to classify 239 patients into the TSSL-PIMS, KDL-PIMS, and U-PIM groups, congruent with the phenotypes identified in previous studies. (13–15) LCA is a probabilistic modeling algorithm that allows clustering cases by associating indicator categorical variables. Three-class LCA was conducted using the R software package “poLCA” with 100 iterations to identify the clusters. The fit of each model was assessed using the Bayesian information criterion score. Indicator variables used in the final LCA model were the 20 features of Kawasaki disease, KDL-MIS-C, NS-MIS-C-TS, and TSSL-MIS-C. (Table 1, Suppl Chart 1).

**Tabl.1.** Clinical presentation of cases with MIS-C.

| Tabl.1. Clinical presentation of cases with MIS-C |  |  |  |  |  |
| --- | --- | --- | --- | --- | --- |
|  | Total population<br>(N=239)<sup>1</sup> | Group 1<br>Toxic shock<br>syndrome like<br>(N=61) | Group 2<br>Kawasaki like<br>(N=70) | Group 3<br>Nonspecific<br>(N=108) | P value<sup>2</sup> |
| Female sex -no. (%) | 103 (43.1%) | 29 (47.5%) | 30 (43.5%) | 44 (40.7%) | 0.68 |
| Age median (IQR) | 7.8(2.6-13.2) | 11.0 (7.1-14.4) | 6.8 (2.4-10.5) | 6.2 (1.9-13.4) | <0.001 |
| <1 y | 30 (12.6%) | 2 (6.7%) | 6 (20%) | 22 (73.3%) |  |
| 1-4 y | 57 (23.9%) | 7 (12.3%) | 26 (45.6%) | 24 (42.1%) |  |
| 5-9 y | 54 (22.6%) | 14 (25.9%) | 19 (35.2%) | 21 (38.9%) |  |
| 10-14 y | 67 (28.03%) | 28 (41.8%) | 17 (25.4%) | 22 (32.8%) |  |
| 15-18 y | 31 (13.0%) | 10 (32.3%) | 2 (6.5%) | 19 (61.3%) |  |
| Days with symptoms before admission | 6 (4-9) | 7 (4-8) | 4 (2-6) | 7 (4-10) | <0.001 |
| **SARS-CoV2 infection evidence** |  |  |  |  |  |
| Positive RT- PCR test | 105/182 (57.7%) | 40/57 (70%) | 16/54 (29.6%) | 49/71 (69%) | <0.001 |
| Positive rapid antigen test | 46/129 (35.7%) | 14/34 (41.2%) | 9/44 (20%) | 23/51 (45.1%) | 0.03 |
| Positive RT-PCR or rapid antigen test | 130/213 (61.0%) | 47/60 (78%) | 23/65 (35.4%) | 60/88 (68.2%) | <0.001 |
| Positive serology | 114/160 (71.3%) | 24/37 (64.9%) | 43/52 (82.7%) | 47/71 (66.2%) | 0.08 |
| **Previous comorbidities** |  |  |  |  |  |
| Obesity (BMI > 95th percentile) | 38 (15.9%) | 5 (8.6%) | 12 (17.6%) | 21 (19.4%) | 0.17 |
| Respiratory | 11 (4.6%) | 3 (6.7%) | 2 (10.5%) | 6 (9.8%) | 0.76 |
| Neuromuscular | 14 (15.9%) | 5 (11.1%) | 3 (15.8%) | 6 (9.8%) | 0.69 |
| Cardiovascular | 6 (2.5%) | 1(2.2%) | 0 | 5(8.2%) | 0.25 |
| Cancer | 16 (6.7%) | 7 (15.6%) | 0 | 9(14.8%) | 0.18 |
| Immunosuppression | 3 (1.3%) | 2 (4.4%) | 0 | 1(1.6%) | 0.74 |
| At least one underlying condition | 83 (34.7%) | 24 (39.3%) | 17 (24.3%) | 42 (38.9%) | 0.09 |
| Severe underlying condition | 41 (17.5%) | 15 (24.6%) | 4 (5.7%) | 22 (20.4%) | 0.004 |
| **Clinical presentation** |  |  |  |  |  |
| Rash | 98 (41%) | 30 (49.2%) | 49 (70%) | 19 (17.6%) | <0.001 |
| Conjunctivitis | 118 (49.4%) | 32 (52.5%) | 59 (84.3%) | 27 (25%) | <0.001 |
| Diarrhea | 80 (33.5%) | 25 (41.0%) | 23 (32.9%) | 32 (29.6%) | 0.31 |
| Emesis | 86 (36.0%) | 29 (47.5%) | 29 (41.4%) | 28 (25.9%) | 0.01 |
| Abdominal pain | 91 (38.1%) | 43 (70.5%) | 33 (47.1%) | 15 (13.9%) | <0.001 |
| Rhinorrhea | 57(23.8%) | 22 (36.1%) | 21 (30%) | 14 (13%) | 0.001 |
| Respiratory dysfunction | 150 (62.8%) | 54 (88.5%) | 25 (35.7%) | 71 (65.7%) | <0.001 |
| Anosmia/Dysgeusia | 11 (4.6%) | 5 (8.2%) | 2 (2.9%) | 4(3.7%) | 0.33 |
| Arthralgias/Myalgias | 48 (20.1%) | 13 (21.3%) | 22 (31.4%) | 13 (12.0%) | 0.007 |
| **Laboratory findings** |  |  |  |  |  |
| Neutrophil count x10<sup>6</sup> cells/mcl | 8.1 (4.2-13.6) | 6.5 (1.7-12.2) | 9.5 (6.3-14.9) | 7.7 (3.7-12.5) | 0.02 |
| Neutrophilia (>7500 cells/mcl) | 130/238 (54.6%) | 30 (49.2%) | 46/69 (66.7%) | 54 (50%) | 0.06 |
| Neutropenia (<1000 cells/mcl) | 22 /239 (9.2%) | 7 (11.5%) | 2 (2.9%) | 13 (12%) | 0.07 |
| Lymphocyte count x10<sup>6</sup> cells/mcl | 1.4 (0.6-2.6) | 0.8 (0.4-1.7) | 1.3 (0.9-2.2) | 1.8 (0.9-3.4) | <0.001 |
| Lymphopenia<sup>a</sup> | 165/237 (69.6%) | 44 (73.3%) | 56/69 (81.2%) | 65 (60.2%) | 0.01 |
| Platelets x 10<sup>3</sup> cells/mcl | 173 (96-315) | 104 (64-165) | 193 (99-329) | 216(148-365) | <0.001 |
| Platelets < 150 x 10<sup>3</sup> cells/mcl | 93/239 (38.9%) | 41 (67.2%) | 25 (35.7%) | 27 (25%) | <0.001 |
| Fibrinogen, mg/dl | 427 (298-559) | 508(348-617) | 401(306-549) | 391 (256-531) | 0.24 |
| Fibrinogen> 400 mg/dl | 104/183 (56.8%) | 39/58 (67.2%) | 24/51 (47.1%) | 41/74 (55.4%) | 0.10 |
| Ferritin, mcg/L | 557 (304-1209) | 828 (508-1956) | 437 (213-793) | 530 (252-1209) | <0.001 |
| Ferritin> 300 mg/dl | 144/191 (75.4%) | 50/54 (92.6%) | 39/59 (66.1%) | 55/78 (70.5%) | 0.001 |
| Ferritin >1500 mg/dl | 32/191 (16.7%) | 14/54 (25.9%) | 2/59 (3.4%) | 16/78 (20.5%) | 0.001 |
| D-Dimer ng/ml | 2.2 (1-5) | 4.3 (1.7-9.2) | 2.1 (1.0-3.8) | 1.6(0.8-4.0) | <0.001 |
| D- Dimer> 560ng/ml (0.56 ng/L) | 194/219 (88.6%) | 55/58 (94.8%) | 57/67 (85.1%) | 82/94 (87.2%) | 0.178 |
| C-reactive protein (mg/L) | 60 (17-180) | 15 (7-31.5) | 38.7 (14.6-179.5) | 22.5 (7.3-86.9) | 0.85 |
| C-reactive protein > 5mg/L | 200/225 (88.9%) | 50/56 (89.3%) | 62/68 (91.2%) | 88/101 (87.1%) | 0.71 |
| Procalcitonin ng/ml | 0.85 (0.14-3.6) | 1.9 (0.5-4.8) | 0.5 (0.0-2.5) | 0.5 (0.1-2.5) | 0.005 |
| Procalcitonin > 0.15ng/ml | 126 /171 (73.7%) | 49/55 (89.1%) | 27/41 (65.9%) | 50/75 (66.7%) | 0.005 |
| Albumin< 3g/dl | 133/222 (59.9%) | 51/61 (83.6%) | 40/66 (60.6%) | 42/95 (44.2%) | <0.001 |
| <sup>1</sup>N in the column heading was used to calculate proportions if not otherwise specified. For variables with missing data, denominators are reported in each cell. |  |  |  |  |  |
| <sup>2</sup> One-sided Fisher exact test/ Mann-Whitney test for comparison between groups 1, 2, and 3. <sup>a</sup>Lymphopenia: <2 years: <4000, 2-3 years: < 3000, >4 years: <1500 cells/mcl |  |  |  |  |  |

Potential heterogeneity among the participating centers was addressed by determining 95% confidence intervals (CIs) of the frequencies of measured outcomes using random-effects meta-analysis. Odds ratios (ORs) with 95% CIs of pooled data were calculated using clustered-robust standard error adjustment. ORs adjusted for the participating centers were calculated using multilevel mixed-effects logistic regression.

## 3. Results

Of the 264 identified patients, 6, 11, and 8 were excluded due to duplicate records, lack of evidence on multi-organ involvement, and lack of evidence of previous SARS-CoV-2 infection, respectively. **Tables 1** and **2** summarize the clinical characteristics and outcomes of the remaining 239 patients included in the final analysis.

LCA indicated that 61 (26%), 70 (29%), and 108 (45%) of the patients exhibited the clinical profiles of TSSL-MIS-C, KDL-MIS-C, and NS-MIS-C, respectively (**Table 1**). The median age was higher in the TSSL-MIS-C group than in the overall cohort (11[IQR:7.1-14.4] versus 7.8[IQR:2.6-13.2] years). Duration of clinical symptoms before diagnosis was significantly shorter in the KDL-MIS-C group than in the other two clinical groups (p < 0.001). Additionally, the frequency of virologic positivity for acute SARS-CoV-2 infection was lower (p <0.001) and the frequency of serologic positivity tended to be higher (p = 0.08) in the KDL-MIS-C group than in the other two clinical groups. Only four (5.7%) children in the KDL-MIS-C group had a severe underlying condition, while it was present in 15(24.6%) and 22(20.4%) of the TSSL-MIS-C and NS-MIS-C, respectively. The levels of plasma inflammatory markers were highest in the TSSL-MIS-C group compared with the other two clinical groups.

**Table 2** describes treatments and outcomes in the total study population and the three clinical groups. IVIG and systemic glucocorticoids were administered in 172 (42%) and 187 (78.2%) of the patients, respectively; both treatments were more frequently (93% and 91%) administered to patients in the KDL-MIS-C group. Anticoagulant therapy was prescribed in 99 (41.4%) of the whole cohort and administered more frequently (60%) in the TSSL-MIS-C group. Oxygen supplementation and vasopressor support were required in 143 (59%) and 92 (39%) of the whole cohort, respectively. Left ventricular ejection fraction was below 55% in 31% of the patients, and coronary artery dilatation and aneurysm were observed in 9 (5.7%) and 21(13.2%) of the patients in whom an echocardiogram was performed, respectively. The median in-hospital stay for the whole cohort was eight days (IQR: 5-13), and about half (54.4%) of the patients were admitted to the ICU. There were 13 in-hospital deaths (5.4%), including eight patients with severe underlying conditions and two who died from causes unrelated to SARS-COV-2 infection. Most deaths (69%) occurred in the TSSL-MIS-C group, whereas no deaths occurred in the KDL-MIS-C group.

**Table 2.**
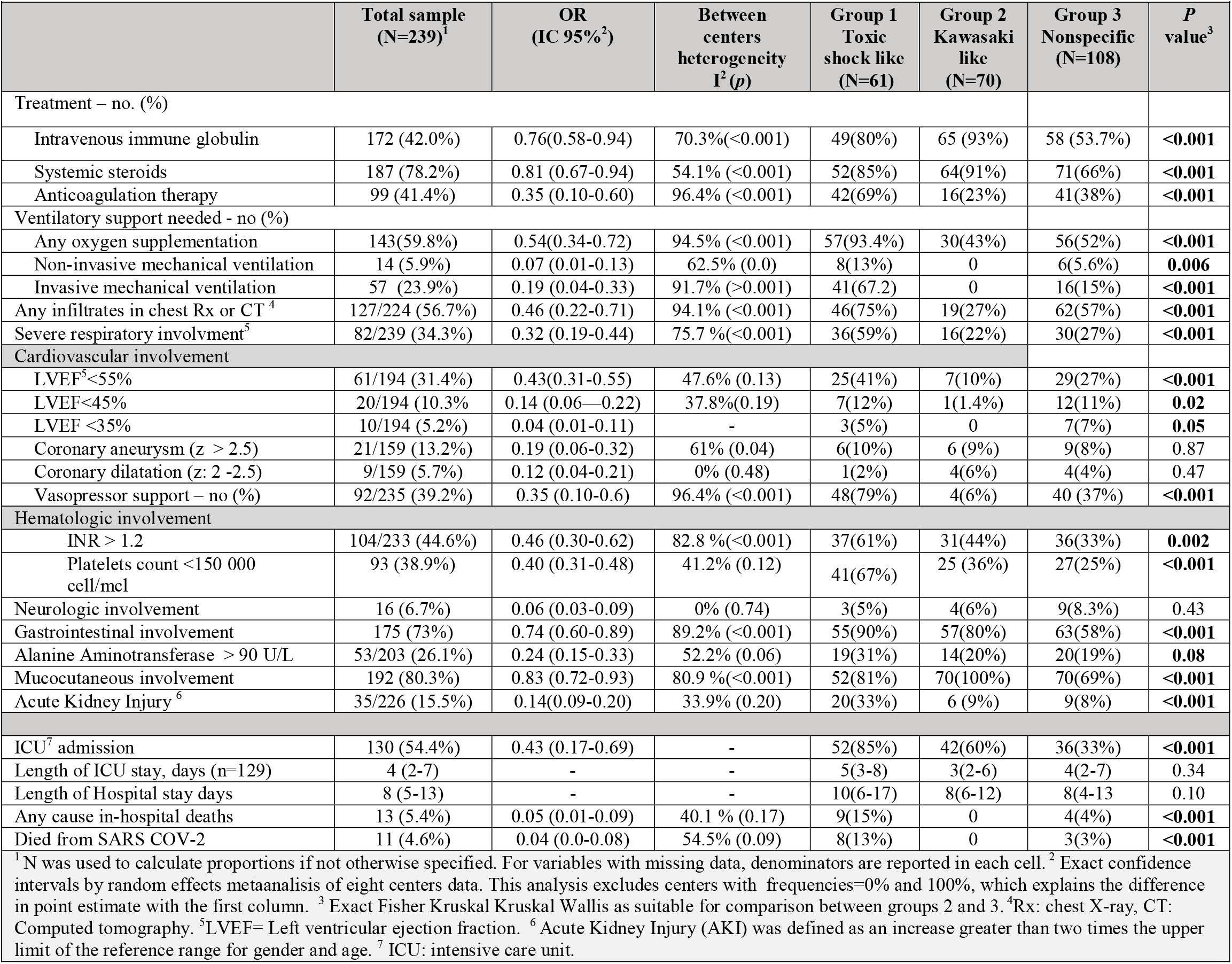
Treatment and outcomes of cases with MIS-C.

Hematologic, neurologic, gastrointestinal, mucocutaneous, and renal dysfunction were frequently observed, with variability in their frequencies among the clinical groups. Prolonged coagulation time based on international normalized ratio, low platelet count, gastrointestinal symptoms, and acute renal injury were more frequent in the TSSL-MIS-C group (61%, 67%, 90%, and 33% of the patients, respectively) than in the other clinical groups.

Coronary aneurysm or dilatation occurred in 27 (11.3%) patients. (2) After adjustment for cluster effect, symptom duration > 10 days before hospital admission was the only factor significantly associated with coronary artery abnormalities (OR 1.6, 95% CI 1.2–2.0). (**Table 3**).

**Table 3.** Coronary artery aneurism or dilatation associated factors.

| **Table 3. Coronary artery aneurism or dilatation associated factors.** |  |  |  |  |  |
| --- | --- | --- | --- | --- | --- |
|  | **Frequency in the aneurism/ dilatation group**<br>**N=27 (%)** | **Unadjusted OR (95% CI) <sup>1</sup>** | ***p*-value** | **OR adjusted for center <sup>2</sup>** | **p-value** |
| Sex (male) | 19/27 (70.4%) | 2.20 (0.99-4.9) | 0.05 | 1.71 (0.63-4.58) | 0.29 |
| Age≥10 years | 5/ 27(18.5%) | 0.31 (0.05-2.04) | 0.22 | 0.55 (0.17-1.80) | 0.32 |
| ≥10 days with symptoms before admission | 10/25 (40%) | **4.16 (2.61-6.23)** | **<0.001** | **1.62 (1.19-2.04)** | **<0.001** |
| Positive SARS-COV-2 RT-PCR or rapid antigen test | 12/20 (60%) | 0.70 (0.13-3.73) | 0.67 | 1.11 (0.31-3.96) | 0.87 |
| Positive SARS-COV-2 serology | 16/20 (80%) | 2.58 (0.77-8.64) | 0.125 | 1.45 (0.36-5.89) | 0.60 |
| Obesity (BMI > 95th percentile) | 16/20 (80%) | 1.04 (0.53-2.04) | 0.92 | 1.40 (0.37-5.37) | 0.63 |
| Rash | 9/27 (33%) | 0.49 (0.10-2.29) | 0.36 | 0.69 (0.25-1.91) | 0.48 |
| Conjunctivitis | 17/27 (63.0%) | 1.5 (0.33-6.92) | 0.60 | 1.82 (0.57-5.77) | 0.31 |
| Gastrointestinal | 16/27 (59.3%) | 0.39 (0.10-1.49) | 0.17 | 0.70 (0.22-2.15) | 0.54 |
| Severe respiratory dysfunction | 11/27 (40.74%) | 1.53 (0.63-3.71) | 0.35 | 1.56 (0.59-4.18) | 0.37 |
| Arthralgias/Myalgias | 2/27 (4.41%) | **0.21 (0.05-0.94)** | **0.04** | 0.13 (0.02-0.67) | 0.015 |
| Mucocutaneous involvement | 13/27 (48.15%) | **0.35 (0.17-0.70)** | **0.003** | 0.74 (0.25-2.17) | 0.59 |
| Neutrophilia (>7500 cells/mcl) | 19/27 (70.37%) | 2.13 (0.82-5.58) | 0.12 | 1.87 (0.67-5.22) | 0.23 |
| Lymphopenia<sup>3</sup> | 15/27 (55.56%) | 0.52 (0.20-1.31) | 0.17 | 0.40 (0.15-1.10) | 0.08 |
| Platelets count <150 000 cell/mcl | 7/27 (25.93%) | 0.48 (0.17-1.40) | 0.18 | 0.48 (0.16-1.37) | 0.17 |
| Fibrinogen> 400 mg/dl | 11/21 (52.38%) | 0.68 (0.26-1.75) | 0.42 | 0.86 (0.31-2.37) | 0.77 |
| Ferritin> 300 mg/dl | 18/20 (90%) | 4.39 (0.81-23.89) | 0.09 | 4.50 (0.90-22.5) | 0.07 |
| D- Dimer> 560ng/ml (0.56 ng/L) | 24/26 (92.3%) | 1.64 (0.41-6.47) | 0.48 | 0.97 (0.17-5.66) | 0.98 |
| C-reactive protein > 5mg/L | 26/27 (96.3%) | 2.41 (0.47-12.4) | 0.29 | 2.68 (0.28-25.31) | 0.39 |
| Procalcitonin > 0.15ng/ml | 15/23 (65.2%) | **0.45 (0.27-0.76)** | **0.003** | 0.84 (0.23-3.00) | 0.75 |
| Albumin< 3g/dl | 20/27 (74.07%) | 1.83 (0.72-4.62) | 0.20 | 1.62 (0.57-4.63) | 0.90 |
| INR > 1.2 | 14/27 (51.85%) | 1.37 (0.54-3.54) | 0.50 | 2.45 (0.86-7.00) | 0.093 |
| LVEF<sup>4</sup><55% | 8 /25 (32%) | 1.25 (0.30-5.1) | 0.76 | 1.49 (0.50-4.47) | 0.45 |
| Alanine Aminotransferase > 90 U/L | 4/25 (16%) | 0.53 (0.19-1.43) | 0.21 | 0.97 (0.26-3.70) | 0.97 |
| Intravenous immune globulin | 27/27 (100%) | - | - | - | - |
| Systemic steroids | 21/27 ( 77.78%) | 0.9 (0.16-4.93) | 0.90 | 0.96 (0.27-3.3) | 0.95 |
| Group 1((TSS-like) | 7/27 (25.9%) | 1 (reference) | - | 1 (reference) | - |
| Group 2 (Kawasaki-like) | 9/27 (33.3%) | 1.14 (0.29-4.45) | 0.85 | 0.66 (0.17-2.58) | 0.55 |
| Group 3 (Unespecific presentation) | 11/27 (40.7%) | 1.25 (0.25-6.15) | 0.78 | 0.59 (0.15-2.31) | 0.45 |
| <sup>1</sup>Clustered robust standard error adjustment. <sup>2</sup> OR adjusted for center with multilevel mixed-effects logistic regression. |  |  |  |  |  |
| <sup>3</sup>Lymphopenia: <2 years: <4000, 2-3 years: < 3000, >4 years: <1500 cells/mcl. <sup>4</sup>LVEF=Left ventricular ejection fraction. |  |  |  |  |  |

Risk factors for death were age ≥ 10 years (OR 5.6, 95% CI 1.4–2.04), presence of at least one underlying condition (OR 5.3, 95% CI 1.5–18.4), severe underlying condition such as cancer (OR 9.3, 95% CI 2.8–31.0), platelet count < 150,000/mm^3^ (OR 4.2, 95% CI 1.2–14.7), international normalized ratio > 1.2 at admission (OR 3.8, 95% CI 1.05–13.9), and serum ferritin concentration > 1500 mg/dL (OR 52, 95% CI 5.9–463) (**Table 4**).

**Table 4.** Characteristics of deaths and associated conditions.

|  | Deaths<br>(N=13) | p<sup>1</sup> | OR (IC95%)<sup>2</sup> |
| --- | --- | --- | --- |
| Female sex -no. (%) | 4 | 0.26 | 0.57 (0.17-1.96) |
| Age-median (IQR) | 14.4 (10.6-15.3) | **0.02** | - |
| <1 y | 2 | - | - |
| 1-4 y | 1 |  |  |
| 5-9 y | 0 |  |  |
| 10-14 y | 6 |  |  |
| 15-18 y | 4 |  |  |
| Age≥10 years | 10 | **0.008** | **5.64 (1.43-22.22)** |
| Days of symptoms before admission | 4 (2-5) | 0.4 |  |
| More than 10 days since symptom onset | 3 | 0.39 | 1.53 (0.36-6.47) |
| Positive RT- PCR test | 8/10 | 0.11 | 3.13 (0.76-12.81) |
| Positive rapid antigen test | 2/6 | 0.49 | 0.80 (0.17-3.69) |
| Positive RT-PCR or rapid antigen test | 8/12 | 0.47 | 1.42 (0.38-5.28) |
| Positive SARS-CoV-2 serology | 3/4 | 0.67 | 0.81 (0.05-13.62) |
| Obesity (BMI > 95th percentile) | 0 | 0.09 | - |
| Respiratory | 1 | 0.58 | 1.08 (0.11-10.82) |
| Neuromuscular | 3 | 0.06 | 3.73 (0.73-19.19) |
| Cardiovascular | 0 | 0.63 | - |
| Cancer | 4 | **0.016** | **6.3 (1.3-30.6)** |
| Immunosuppression | 1 | 0.20 | 6.99 (0.41-119.11) |
| End-stage chronic renal disease | 1 |  | 1.69 (0.15-18.7) |
| At least one underlying condition | 9 | **0.01** | **5.3 (1.5-18.4)** |
| Severe underlying condition | 8 | **<0.001** | **9.3 (2.8-31.0)** |
| On palliative care | 1 | - | - |
| Lymphocyte count x10<sup>6</sup> cells/mcl -median (IQR) | 0.6 (0.3-1.4) | **0.01** | - |
| Lymphopenia<sup>a</sup> | 10 | 0.40 | 1.4 (0.4-5.5) |
| Platelets x 10<sup>3</sup> cells/mcl | 94 (46-191) | **0.03** | - |
| Platelets < 150 x 10<sup>3</sup> cells/mcl _median (IQR) | 9 | **0.023** | **4.2 (1.2-14.7)** |
| Fibrinogen, mg/dl | 94 (46-191) | 0.89 | - |
| Fibrinogen> 400 mg/dl | 6/8 | 0.24 | 2.48 (0.47-13.15) |
| Ferritin, mcg/-L median (IQR) | 3619 (2108-4752) | **<0.001** | - |
| Ferritin >300 | 9/9 | **0.04** | - |
| Ferritin >1500 | 8/9 | **<0.001** | **52 (5.9-463)** |
| Intravenous immune globulin | 8 | 0.28 | 0.34 (0.08-1.44) |
| Systemic steroids | 11 | 0.43 | 1.28 (0.25-6.61) |
| Anticoagulation therapy | 7 | 0.25 | 1.52 (0.45- 5.15) |
| Coronary aneurysm (z score > 2.5) | 0/2 | 0.75 | - |
| Coronary dilatation (z score >2 -<2.5) | 0/2 | 0.89 | - |
| INR > 1.2 | 9 | **0.06** | **3.8 (1.05-13.9)** |
| ICU<sup>3</sup> admission | 11 | 0.02 | 6.5 (1.14-37.11) |
| Group 1(TSS-like) | 9 | **0.001** | 1 (reference) |
| Group 2) (Kawasaki like) | 0 |  | - |
| Group 3 (Unespecific presentation) | 4 |  | **0.15 (0.03-0.66)** |

1 Comparison with surviving cases with one tailed exact Fisher test or Mann-Whitney U test. <sup>2</sup>OR adjusted by center with multilevel mixed effects logistic regression. <sup>3</sup>ICU= Intensive care unit. <sup>a</sup>Lymphopenia: <2 years: <4000, 2-3 years: < 3000, >4 years: <1500 cells/mcl

The patient population was not homogeneous among the eight participating centers. Heterogeneity was high for almost all calculated treatment and outcome frequencies, and I^2^, a measure of heterogeneity, was 50% for most of the factors. Accordingly, association measures were adjusted for the center effect. A significant cluster effect of the reporting center was demonstrated for every evaluated factor in **Tables 3** and **4**. Disaggregated data collected from the five centers with more included cases are presented in **Supplementary Tables 1** and **2**.

## 4. Discussion

MIS-C, a recently characterized clinical presentation secondary to SARS-CoV-2 infection, is closely associated with Kawasaki disease and toxic shock syndrome. In the present study, we aimed to summarize the clinical characteristics and outcomes of patients with MIS-C treated in eight pediatric tertiary care centers across Mexico. Our analyses reveal that these patients share several similarities with previous case series while exhibiting some differences and illustrate the presence of high heterogeneity in disease presentation and outcomes among the participating centers.

It is relevant and essential to examine the clinical characteristics, disease management, and outcomes of MIS-C due to the potentially severe consequences such as death (16–18), damage to coronary arteries, and the toll on healthcare resources necessary for its management. The present cohort’s mortality rate was 4.6%, which exhibited high heterogeneity across the centers, ranging from 0% to 13.5%. Similarly, the rate of coronary artery abnormalities, which was 18.9% in the overall cohort, ranged from 0% to 56.3% across the participating centers. Published systematic reviews estimate mortality and a rate of coronary artery abnormalities ranging from 0% to 4% and from 11.6% to 21.7%, respectively, in patients with MIS-C, according to the systematic reviews (19–23). In the present study, coronary artery abnormalities were observed in all three clinical groups, with no significant difference in frequency among the groups.

Among the factors that might explain the high heterogeneity in outcomes between the centers included in our study and the previously published case series are (i) a wide spectrum of clinical presentation in the presence of relatively nonspecific diagnostic criteria that are currently used for MIS-C, (ii) variations in ethnicity and prevalence of comorbid conditions, and (iii) variations in access, diagnostic paths, and therapeutic interventions across different healthcare systems.

Attempts have been made to classify cases according to their predominant clinical characteristics to address the lack of specific diagnostic criteria and, thus, phenotype heterogeneity. In the present study, we utilized LCA to classify 239 patients into the TSSL-MIS-C, KDL-MIS-C, and U-PIM groups, congruent with the phenotypes identified in previous studies (13–15) In the present study, patient age was different among the clinical groups; additionally, the frequency of virologic positivity was lower, and the frequency of serologic positivity was higher in the KDL-MIS-C group than in the other two clinical groups, suggesting different underlying pathologic mechanisms at play. Multiple pathologic mechanisms have been shown to be altered in MIS-C, including the possible involvement of superantigens and autoantibodies. (24) The levels of plasma inflammatory markers were highest in the TSSL-MIS-C group. Additionally, the rate of life support measures and mortality rate were highest in this group, as expected. The vagueness of the current clinical criteria for MIS-C might be partially responsible for the wide variation in the rate of these three clinical phenotypes reported by each center, suggesting a lack of clarity among physicians for the threshold of clinical manifestations that eventually leads to the diagnosis of MIS-C (**Supplementary Table 1**).

Demographic and preexisting medical conditions of patients in different published series and clinical groups widely vary, likely accounting for some of the heterogeneity observed in outcomes.

In the present study, 34.7% of the cohort had underlying conditions, which was higher than that reported by Radia et al. and Antúnez et al. (25) In the present cohort, severe underlying conditions were present in 17.5% of the patients varying from 3.6% to 23.1% across the participating centers. In addition, the frequency of patients with cancer/immunosuppressive disorders or neuromuscular conditions was four to tenfold. In contrast, the observed frequency of obesity (15.9%) was not as high as that reported by other studies. (26) Notably, 9 of the 13 deaths in the present cohort had a severe underlying condition, and the mortality was highest in the center with the highest rate of patients with severe underlying conditions and the highest proportion of patients with TSSL-MIS-C. These findings suggest that the high rate of severe underlying conditions might explain the relatively high mortality observed in the present cohort.

Although the centers included in the present study are tertiary care reference hospitals, they belong to different healthcare systems with different protocols for service delivery and different resources for the management of patients during the COVID-19 pandemic; the difference in frequencies of IVIG utilization (from 45% to 94%) might be a reflection of this aspect. In addition, there is evidence of an association between delayed access to health services and worst outcomes, which might explain differences in risk for bad outcomes across different geographic locations(27)Of note, the only factor that was significantly associated with the development of coronary artery abnormalities after adjustment for center effect was a delay of ≥10 days in hospital admission after the onset of symptoms, a factor likely influenced by socioeconomic and geographic factors.

Factors associated with mortality were age ≥ 10 years, severe underlying condition, thrombocytopenia, hyperferritinemia, and toxic shock syndrome phenotype. A study in a larger cohort might reveal an association with other factors, such as IVIG use and coagulopathy. Abrams et al. reported that older age, non-Hispanic black ethnicity, disease onset before June 1, 2020, and high levels of serum D-dimer, troponin, atrial natriuretic peptide, C-reactive protein, ferritin, and interleukin-6 were associated with ICU admission in patients with MIS-C. The authors also reported abdominal pain and shortness of breath as clinical findings at hospitalization admission predicting subsequent ICU admission. (28)

According to most systematic reviews, myocarditis is the most frequent complication observed in patients with MIS-C; however, it was not examined in the present study due to the lack of a standardized definition among the participating centers. However, left ventricular ejection fraction, which was below 55% in 31% of the cohort, might be considered a proxy for this outcome. In addition, we did not evaluate socioeconomic status, ethnicity, and the levels of D-dimer, troponin-B, natriuretic peptide, or interleukin-6, which were assessed in previous studies.

In conclusion, death was an infrequent outcome in patients with MIS-C, occurring mainly in older patients with severe underlying conditions, whereas coronary aneurysms were primarily associated with delayed treatment. Extrapolation of the frequency of observed outcomes to the general population might not be valid due to the high frequency of comorbid conditions and the heterogeneity of participating centers. The causal association of risk factors with adverse outcomes should be further analyzed in longitudinal studies with sufficient statistical power to adjust for multiple confounding factors found in the present cohort.

## Data Availability

Datasets are available on request to NGG

## 6. Author Contributions

MFCP, NGG, and HMG contributed to the conception and design of the study. HMG coordinated interinstitutional collaboration. NGG organized the database. MFCP and NGG performed the statistical analysis. MAYN, NGG, and MFCP wrote the first draft of the manuscript. All authors contributed to data gathering and manuscript revision and read and approved the submitted version.

## 8. Acknowledgments

We acknowledge the authorities of participating centers for the facilities to perform this study.

## 9. Data Availability Statement

Datasets are available on request to NGG

---

**Chart 1.**
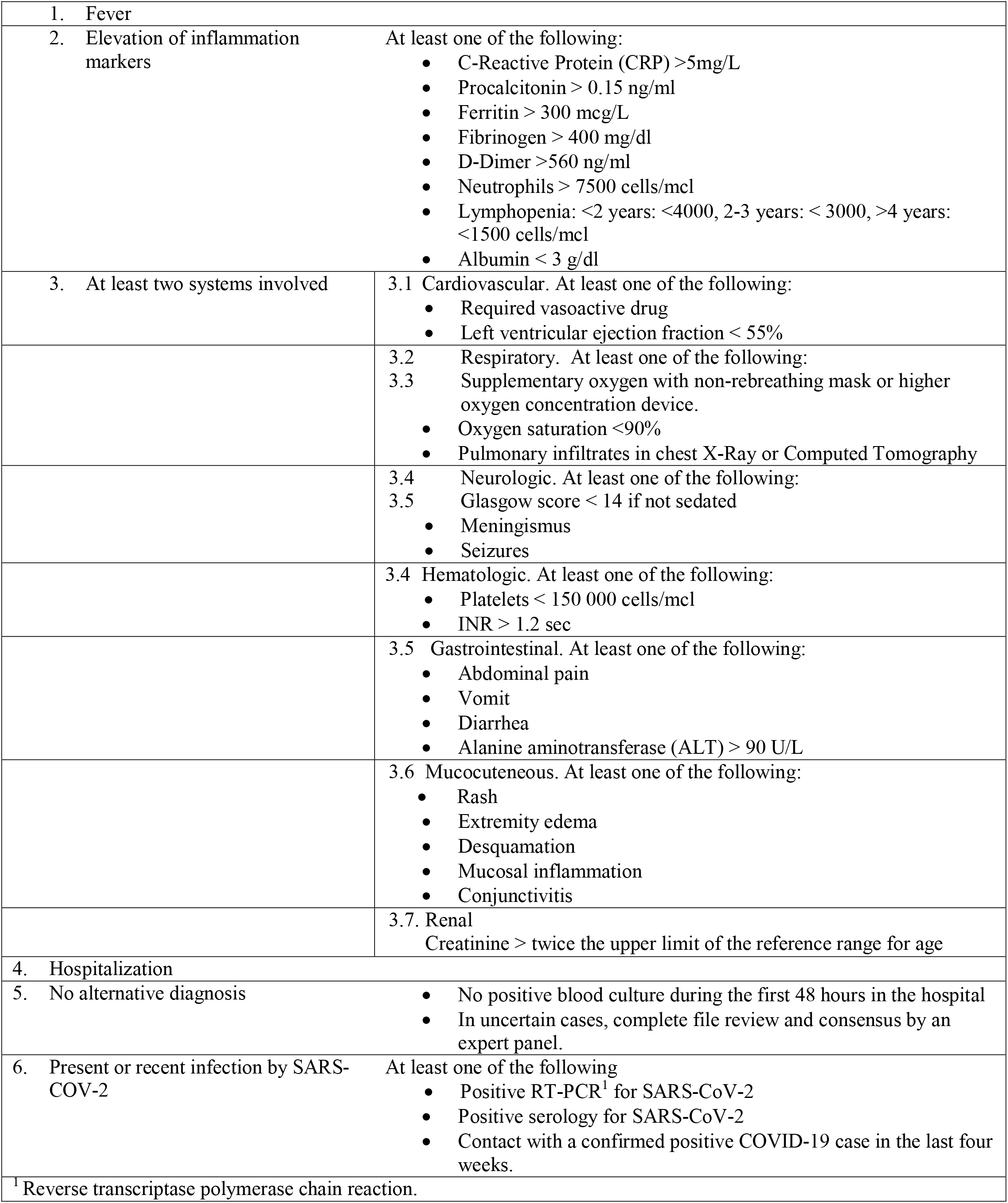
Case definition criteria for MIS-C.

| Chart 1. Case definition criteria for MIS-C |  |
| --- | --- |
| 1. Fever |  |
| 2. Elevation of inflammation markers | At least one of the following:<br>• C-Reactive Protein (CRP) >5mg/L<br>• Procalcitonin > 0.15 ng/ml<br>• Ferritin > 300 mcg/L<br>• Fibrinogen > 400 mg/dl<br>• D-Dimer >560 ng/ml<br>• Neutrophils > 7500 cells/mcl<br>• Lymphopenia: <2 years: <4000, 2-3 years: < 3000, >4 years: <1500 cells/mcl<br>• Albumin < 3 g/dl |
| 3. At least two systems involved | 3.1 Cardiovascular. At least one of the following:<br>• Required vasoactive drug<br>• Left ventricular ejection fraction < 55% |
|  | 3.2 Respiratory. At least one of the following: |
|  | 3.3 Supplementary oxygen with non-rebreathing mask or higher oxygen concentration device.<br>• Oxygen saturation <90%<br>• Pulmonary infiltrates in chest X-Ray or Computed Tomography |
|  | 3.4 Neurologic. At least one of the following: |
|  | 3.5 Glasgow score < 14 if not sedated<br>• Meningismus<br>• Seizures |
|  | 3.4 Hematologic. At least one of the following:<br>• Platelets < 150 000 cells/mcl<br>• INR > 1.2 sec |
|  | 3.5 Gastrointestinal. At least one of the following:<br>• Abdominal pain<br>• Vomit<br>• Diarrhea<br>• Alanine aminotransferase (ALT) > 90 U/L |
|  | 3.6 Mucocutaneous. At least one of the following:<br>• Rash<br>• Extremity edema<br>• Desquamation<br>• Mucosal inflammation<br>• Conjunctivitis |
|  | 3.7. Renal |
|  | Creatinine > twice the upper limit of the reference range for age |
| 4. Hospitalization |  |
| 5. No alternative diagnosis | • No positive blood culture during the first 48 hours in the hospital<br>• In uncertain cases, complete file review and consensus by an expert panel. |
| 6. Present or recent infection by SARS-COV-2 | At least one of the following<br>• Positive RT-PCR<sup>1</sup> for SARS-CoV-2<br>• Positive serology for SARS-CoV-2<br>• Contact with a confirmed positive COVID-19 case in the last four weeks. |
| <sup>1</sup> Reverse transcriptase polymerase chain reaction. |  |

**Table Suppl.1.** Clinical presentation of cases with MIS-C for the five centers with more included cases.

|  | Total population<br>(N=239)<sup>1</sup> | Center 1<br>(N=75) | Center 2<br>(N=52) | Center 3<br>(N=28) | Center 4<br>(N=45) | Center 5<br>(N=30) |
| --- | --- | --- | --- | --- | --- | --- |
| Female sex -no. (%) | 103 (43.1%) | 39 (52%) | 21 (40.4%) | 6 (21.4%) | 17 (38.6%) | 15 (50%) |
| Age - median (IQR) | 7.8(2.6-13.2) | 11.1 (5.7-15.7) | 10.6 (6.6-14.6) | 7.0 (2.6-9.6) | 2.4 (0.7-7.6) | 3.6 (1.3-10.2) |
| <1 y | 30 (12.6%) | 5(6.7%) | 3 (5.8%) | 1 (3.6%) | 15 (13.3%) | 5 (16.7%) |
| 1-4 y | 57 (23.9%) | 11 (14.7%) | 8 (15.4%) | 8 (28.6%) | 14 (31.1%) | 12 (40%) |
| 5-9 y | 54 (22.6%) | 14 (18.7%) | 12 (23.1%) | 13 (46.4%) | 8 (17.8%) | 5 (16.7%) |
| 10-14 y | 67 (28.03%) | 25 (33.3%) | 19 (36.5%) | 6 (21.4%) | 8 (17.8%) | 7 (23.3%) |
| 15-18 y | 31 (13.0%) | 20 (26.7%) | 10 (19.2%) | 0 | 0 | 1 (3.3%) |
| Days with symptoms before admission | 6 (4-9) | 7 (4-7) | 5 (3-9) | 6 (4-7) | 8 (5-14) | 3.5 (2-6) |
| **SARS-COV2 testing No. (%)** |  |  |  |  |  |  |
| Positive RT- PCR test | 105/182 (57.7%) | 57/71 (80.3%) | 13/33 (39.4%) | 5/18 (27.8%) | 17/39 (43.6%) | 6/12 (50%) |
| Positive rapid antigen test | 46/129 (35.7%) | 20/36 (55.6%) | 9/34 (26.5%) | 1/11 (9.1%) | 7/23 (30.4%) | 5/19 (26.3%) |
| Positive RT-PCR or rapid antigen test | 130/213 (61.0%) | 71/75 (94.7%) | 32/52(61.5%) | 4/28 (14.3%) | 11/22 (50%) | 7/27 (25.9%) |
| Positive SARS-COV-2 serology | 114/160 (71.3%) | 25/59 (42.4%) | 16/19 (82.6%) | 18/21 (85.7%) | 32/33 (97.0%) | 19/23 (82.6%) |
| **Underlying medical conditions No. (%)** |  |  |  |  |  |  |
| Obesity (BMI > 95th percentile) | 38 (15.9%) | 14 (18.7%) | 8 (15.4%) | 4 (14.3%) | 4 (8.9%) | 4 (13.3%) |
| Respiratory | 11 (4.6%) | 5 (6.7%) | 2 (3.9%) | 0 | 2 (4.4%) | 1 (3.3%) |
| Neuromuscular | 14 (15.9%) | 6 (8%) | 4 (7.7%) | 1 (3.6 %) | 2 (4.4%) | 1 (3.3%) |
| Cardiovascular | 6 (2.5% ) | 3 (4%) | 2 (3.9%) | 0 | 1 (2.2%) | 0 |
| Cancer | 16 (6.7%) | 7 (9.3%) | 3 (5.8%) | 0 | 4 (8.9%) | 0 |
| Immunosuppression | 3 (1.3%) | 2 (2.7%) | 1 (1.9%) | 0 | 4 (8.9%) | 15 (50%) |
| At least one underlying condition | 83 (34.7%) | 34 (45.3%) | 16 (30.8%) | 1(3.6%) | 15 (33.3%) | 15 (50%) |
| Severe underlying condition | 41 (17.5%) | 16 (21.3%) | 12 (23.1%) | 1 (3.6%) | 8 (17.8%) | 3 (10%) |
| Rash | 98 (41%) | 36 (48%) | 18 (34.6%) | 17(60.7%) | 3 (6.7%) | 20 (66.7%) |
| Conjunctivitis | 118 (49.4%) | 30 (40%) | 39 (75%) | 17 (60.7%) | 6 (13.3%) | 21 (70%) |
| Diarrhea | 80 (33.5%) | 33 (44%) | 11 (21.2%) | 13 (46.4%) | 10 (22.2%) | 12 (40%) |
| Emesis | 86 (36.0%) | 36 (48%) | 15 (28.9%) | 22 (78.6%) | 1 (2.2%) | 11 (36.7%) |
| Abdominal pain | 91 (38.1%) | 38 (50.7%) | 24(46.2%) | 13 (46.4%) | 2 (4.4%) | 12 (40%) |
| Rhinorrhea | 57(23.8%) | 24 (32%) | 25 (48.1%) | 0 | 0 | 8 (26.7%) |
| Respiratory dysfunction | 150 (62.8%) | 67 (89.3%) | 42 (80.8%) | 8 (28.6%) | 21 (46.7%) | 6 (20%) |
| Anosmia/Dysgeusia | 11 (4.6%) | 7 (9.3%) | 3 (5.8%) | 0 | 1 (2.2%) | 0 |
| Arthralgias/Myalgias | 48 (20.1%) | 14 (18.7%) | 17 (32.7%) | 9 (32.1%) | 0 | 7 (23.3%) |
| Respiratory support with high flow cannula or more | 73 (30.5%) | 39 (52%) | 16 (30.1%) | 4 (14.3%) | 10 (22.2%) | 4 (13.3%) |
| Neutrophil count x10<sup>6</sup> cells/mcl | 8.1 (4.2-13.6) | 6.1 (1.7-9.1) | 8.5(5.5-13.3) | 11.1 (6.9-15.9) | 5.6 (4.5-15.5) | 9.7(5.9-15.5) |
| Neutrophilia (>7500 cells/mcl) | 130/238 (54.6%) | 29/74 (39.2%) | 33/52 (63.5%) | 20/28 (71.4%) | 23/45 (51.1%) | 21/30 (70%) |
| Neutropenia (<1000 cells/mcl) | 22 /239 (9.2%) | 11/75 (14.7%) | 3 (5.8%) | 1 (3.6%) | 5 (11.1%) | 1 (3.3%) |
| Lymphocyte count x10<sup>6</sup> cells/mcl | 1.4 (0.6-2.6) | 1.5 (0.6-2.4) | 0.8 (0.5-1.5) | 1.2 (0.7-1.8) | 2.9 (1.6-4.5) | 1.4(1.0-2.1) |
| Lymphopenia<sup>a</sup> | 165/237 (69.6%) | 43/73 (58.9%) | 42 (80.1%) | 25 (89.3%) | 22 (48.9%) | 26 (86.7%) |
| Platelets x 10<sup>3</sup>cells/mcl | 173 (96-315) | 161 (94-236) | 176 (107-249) | 152 (75-386) | 289 (155-533) | 175 (87-340) |
| Platelets < 150 x 10<sup>3</sup>cells/mcl | 93/239 (38.9%) | 30/75 (40%) | 20/52 (38.5%) | 14/28 (50%) | 11/45 (24.4%) | 12/30 (40%) |
| Fibrinogen, mg/dl | 427 (298-559) | 1284 (495-3641) | 452 (326-600) | 404 (313-496) | 250 (197-304) | 313 (226-544) |
| Fibrinogen> 400 mg/dl | 104/183 (56.8%) | 52/75 (69.3%) | 28/50 (56%) | 14/27 (51.9%) | 3/13 (23.08%) | 4/10 (40%) |
| Ferritin, mcg/L | 557 (304-1209) | 618 (410-1340) | 629 (457-1617) | 761 (246-921) | 343 (184-708) | 329 (166-593) |
| Ferritin> 300 mg/dl | 144/202 (71.3%) | 53/75 (70.7%) | 37/44 (84.1%) | 14/19 (73.7%) | 17/28 (60.7%) | 17/29 (58.6%) |
| D-Dimer ng/ml | 2.2 (1-5) | 2.6 (1.1-6.4) | 2.6(1.7-5.4) | 1-6(1.0-3.2) | 1.6(0.6-3.9) | 1.4(0.4-2.4) |
| D- Dimer> 560ng/ml (0.56 ng/L) | 194/219 (88.6%) | 66/74 (89.2%) | 48/49 (98%) | 25/28 (89.3%) | 27/32 (84.4%) | 20/28 (71.4%) |
| C-reactive protein (mg/L) | 60 (17-180) | 100 (43-190) | 13.9 (7.1-23.14) | 168 (112-293) | 33.6 (13.2-190) | 101 (34-209) |
| C-reactive protein > 5mg/L | 200/225 (88.9%) | 58/63 (92.1%) | 39 (76.5%) | 28/28 (100%) | 40/45 (88.9%) | 28/30 (93.3%) |
| Procalcitonin ng/ml | 0.85 (0.14-3.6) | 1.85 (0.5-6.23) | 0.06(0.01-0.53) | 2.24 (0.82-6.0) | 0.40 (0.14-4.5) | 0.5 (1.5-1.5) |
| Procalcitonin > 0.15ng/ml | 126 /171 (73.7%) | 55/62 (88.7%) | 14/36 (38.9%) | 15/16 (93.8%) | 22/32 (68.8%) | 13/17 (76.5%) |
| Albumin< 3g/dl | 133/222 (59.9%) | 45/71 (63.4%) | 25(51.0%) | 19/28 (67.9%) | 23/37 (62.2%) | 16 (55.2%) |
| **Clinical profile** |  |  |  |  |  |  |
| Group 1(Kawasaki like) | 61 (25.5%) | 36 (48%) | 15 (28.9%) | 7 (25%) | 2 (4.4%) | 1 (3.3%) |
| Group 2(TSS-like) | 70 (29.3%) | 2 (2.7%) | 22 (42.3%) | 12 (42.9%) | 2 (4.4%) | 27 (90%) |
| Group 3 (Unespecific presentation) | 108 (45.2%) | 37 (49.3%) | 15 (28.9%) | 9(32.1%) | 41 (91%) | 2 (3.7%) |

<sup>1</sup>N in the column heading was used to calculate proportions if not otherwise specified. For variables with missing data, denominators are reported in each cell, lymphopenia<sup>a</sup>: <2 years: <4000, 2-3 years: < 3000, >4 years: <1500 cells/mcl

**Table Suppl.2.**
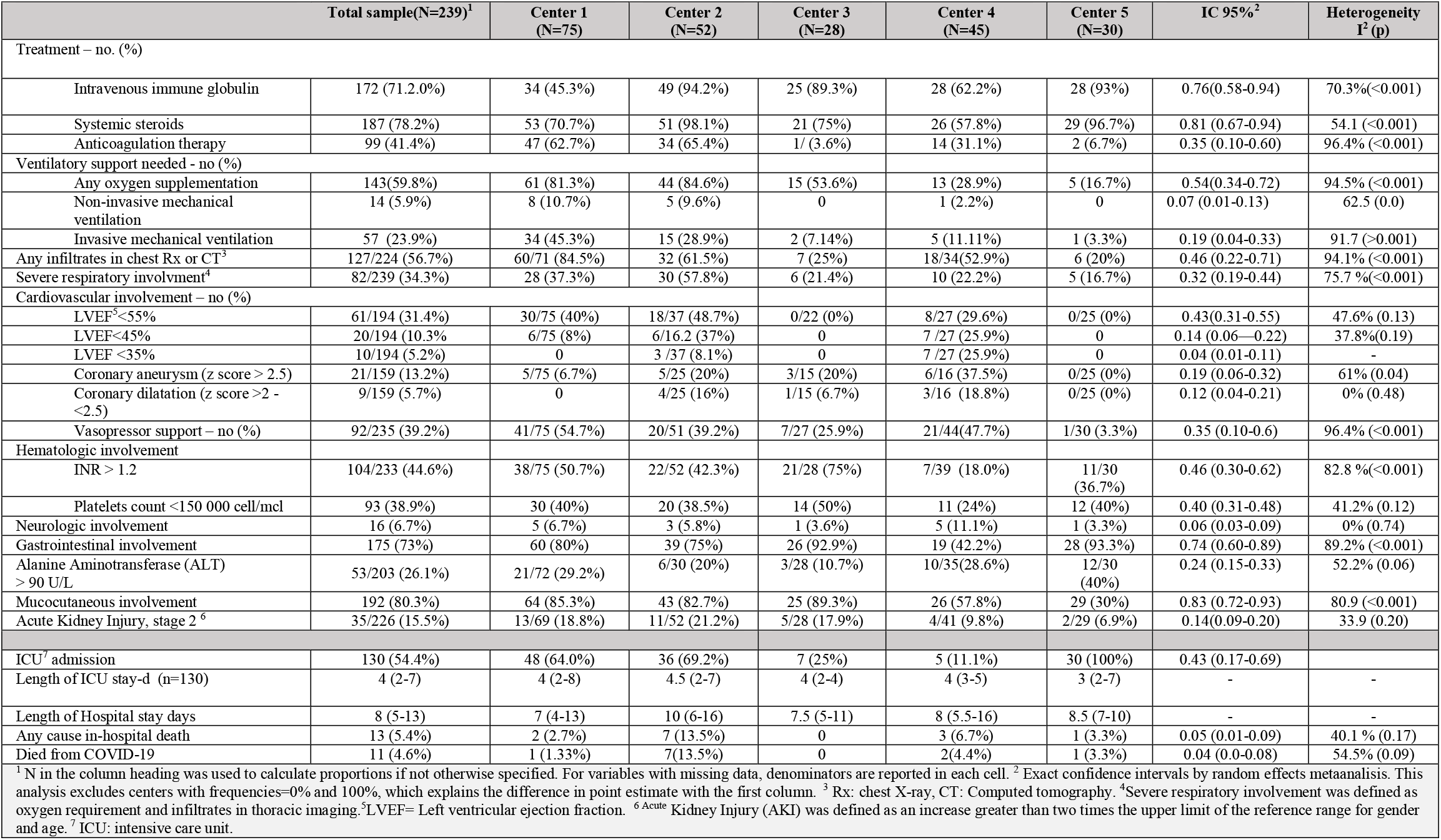
Treatment and outcomes of cases with MIS-C for the five centers with more included cases.

**Supplementary Chart 1.**
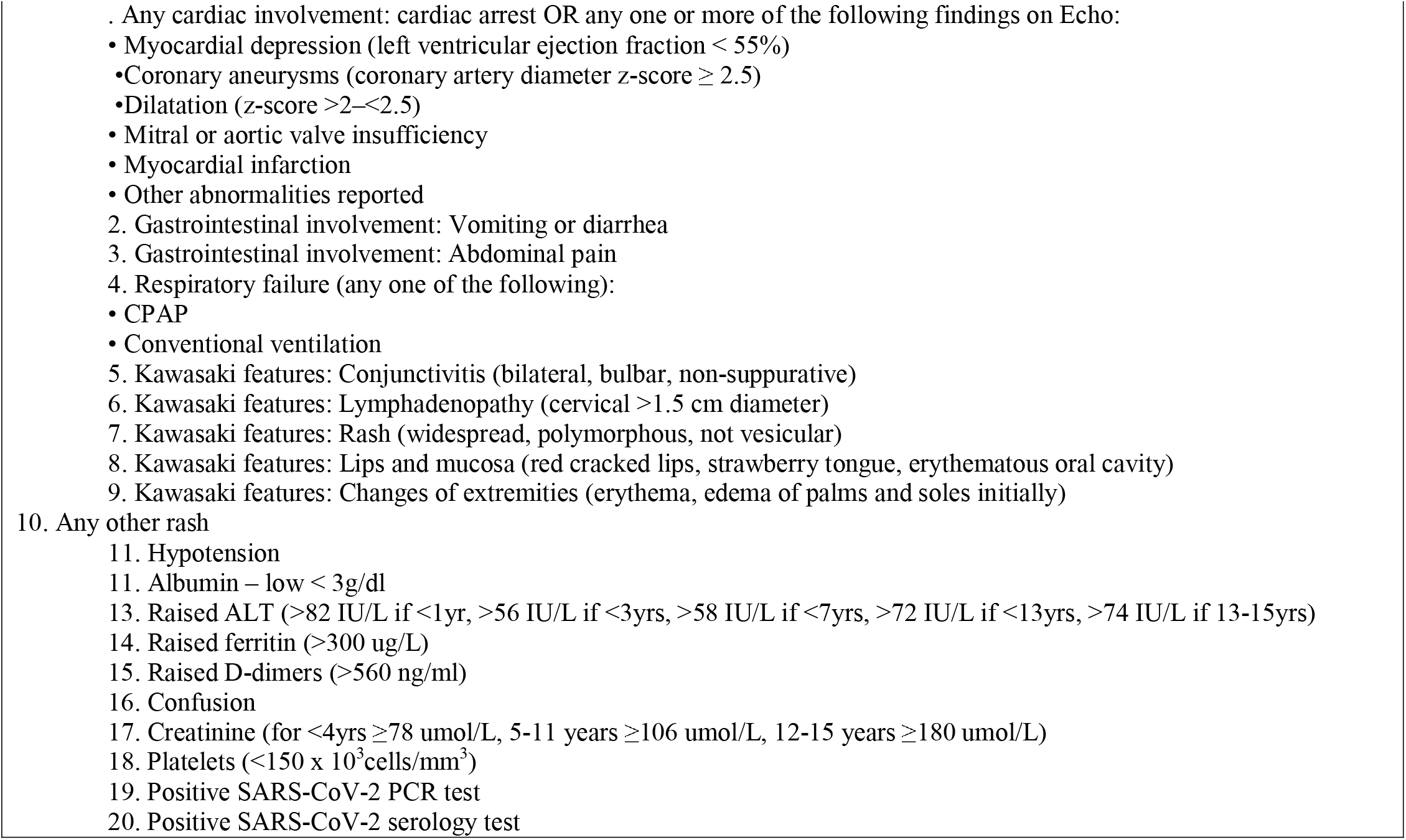
Variables included in Latent Class Analysis.

| Supplementary Chart 1. Variables included in Latent Class Analysis |
| --- |
| Any cardiac involvement: cardiac arrest OR any one or more of the following findings on Echo:<br>Myocardial depression (left ventricular ejection fraction < 55%)<br>Coronary aneurysms (coronary artery diameter z-score ≥ 2.5)<br>Dilatation (z-score >2–<2.5)<br>Mitral or aortic valve insufficiency<br>Myocardial infarction<br>Other abnormalities reported<br>Gastrointestinal involvement: Vomiting or diarrhea<br>Gastrointestinal involvement: Abdominal pain<br>Respiratory failure (any one of the following):<br>CPAP<br>Conventional ventilation<br>Kawasaki features: Conjunctivitis (bilateral, bulbar, non-suppurative)<br>Kawasaki features: Lymphadenopathy (cervical >1.5 cm diameter)<br>Kawasaki features: Rash (widespread, polymorphous, not vesicular)<br>Kawasaki features: Lips and mucosa (red cracked lips, strawberry tongue, erythematous oral cavity)<br>Kawasaki features: Changes of extremities (erythema, edema of palms and soles initially)<br>Any other rash<br>Hypotension<br>Albumin – low < 3g/dl<br>Raised ALT (>82 IU/L if <1yr, >56 IU/L if <3yrs, >58 IU/L if <7yrs, >72 IU/L if <13yrs, >74 IU/L if 13-15yrs)<br>Raised ferritin (>300 ug/L)<br>Raised D-dimers (>560 ng/ml)<br>Confusion<br>Creatinine (for <4yrs ≥78 umol/L, 5-11 years ≥106 umol/L, 12-15 years ≥180 umol/L)<br>Platelets (<150 x 10<sup>3</sup> cells/mm<sup>3</sup>)<br>Positive SARS-CoV-2 PCR test<br>Positive SARS-CoV-2 serology test |

## References

1. Licciardi F, Pruccoli G, Denina M, Parodi E, Taglietto M. SARS-CoV-2 – Induced Kawasaki-Like Hyperin fl ammatory Syndrome□: A Novel COVID Phenotype in Children. Pediatrics. 2021;146(2):1–5.

2. HAN Archive - 00432 Health Alert Network (HAN) [Internet]. [cited 2021 Dec 13]. Available from: https://emergency.cdc.gov/han/2020/han00432.asp

3. Royal College of Paediatrics and Child Health. Guidance paediatric multisystem inflammatory syndrome temporallly associated with Cov-19. Royal College of Paediatrics and Child Health. 2020;1–6.

4. Jiang L, Tang K, Levin M, Irfan O, Morris SK, Wilson K, et al. COVID-19 and multisystem inflammatory syndrome in children and adolescents. Lancet Infect Dis. 2020;20(11):e276–88.

5. Jiang L, Tang K, Levin M, Irfan O, Morris SK, Wilson K, et al. COVID-19 and multisystem inflammatory syndrome in children and adolescents. Lancet Infect Dis. 2020;20(11):e276–88.

6. Feldstein LR, Tenforde MW, Friedman KG, Newhams M, Rose EB, Dapul H, et al. Characteristics and Outcomes of US Children and Adolescents with Multisystem Inflammatory Syndrome in Children (MIS-C) Compared with Severe Acute COVID-19. JAMA - Journal of the American Medical Association. 2021;325(11):1074–87.

7. Son MBF, Murray N, Friedman K, Young CC, Newhams MM, Feldstein LR, et al. Multisystem Inflammatory Syndrome in Children — Initial Therapy and Outcomes. New England Journal of Medicine. 2021;385(1):23–34.

8. Davies P, Evans C, Kanthimathinathan HK, Lillie J, Brierley J, Waters G, et al. Intensive care admissions of children with paediatric inflammatory multisystem syndrome temporally associated with SARS-CoV-2 (PIMS-TS) in the UK: a multicentre observational study. Lancet Child Adolesc Health. 2020;4(9):669–77.

9. Riphagen S, Gomez X, Gonzalez-Martinez C, Wilkinson N, Theocharis P. Hyperinflammatory shock in children during COVID-19 pandemic. The Lancet. 2020;395(10237):1607–8.

10. Dufort EM, Koumans EH, Chow EJ, Rosenthal EM, Muse A, Rowlands J, et al. Multisystem Inflammatory Syndrome in Children in New York State. New England Journal of Medicine. 2020;383(4):347–58.

11. Belay ED, Abrams J, Oster ME, Giovanni J, Pierce T, Meng L, et al. Trends in Geographic and Temporal Distribution of US Children with Multisystem Inflammatory Syndrome during the COVID-19 Pandemic. JAMA Pediatr. 2021;175(8):837–45.

12. Payne AB, Gilani Z, Godfred-Cato S, Belay ED, Feldstein LR, Patel MM, et al. Incidence of Multisystem Inflammatory Syndrome in Children Among US Persons Infected With SARS-CoV-2. JAMA Netw Open [Internet]. 2021 Jun 1 [cited 2021 Dec 13];4(6):e2116420–e2116420. Available from: https://jamanetwork.com/journals/jamanetworkopen/fullarticle/2780861

13. Flood J, Shingleton J, Bennett E, Walker B, Amin-Chowdhury Z, Oligbu G, et al. Paediatric multisystem inflammatory syndrome temporally associated with SARS-CoV-2 (PIMS-TS): Prospective, national surveillance, United Kingdom and Ireland, 2020. The Lancet Regional Health - Europe [Internet]. 2021 Apr 1 [cited 2021 Dec 13];3. Available from: http://www.thelancet.com/article/S2666776221000521/fulltext

14. Godfred-Cato S, Abrams JY, Balachandran N, Jaggi P, Jones K, Rostad CA, et al. Distinguishing Multisystem Inflammatory Syndrome in Children from COVID-19, Kawasaki Disease and Toxic Shock Syndrome. Pediatric Infectious Disease Journal. 2022 Apr 1;41(4):315–23.

15. Geva A, Patel MM, Newhams MM, Young CC, Son MBF, Kong M, et al. Data-driven clustering identifies features distinguishing multisystem inflammatory syndrome from acute COVID-19 in children and adolescents. EClinicalMedicine [Internet]. 2021;40:09–10. Available from: https://doi.org/10.1016/j.eclinm.2021.101112

16. Laverty M, Salvadori M, Squires SG, Ahmed M, Eisenbeis L, Lee S, et al. Multisystem inflammatory syndrome in children in Canada. Canada Communicable Disease Report [Internet]. 2021 Nov 10;47(11):461–5. Available from: https://www.canada.ca/content/dam/phac-aspc/documents/services/reports-publications/canada-communicable-disease-report-ccdr/monthly-issue/2021-47/issue-11-november-2021/ccdrv47i11a03-eng.pdf

17. Baradaran A, Malek A, Moazzen N, Shaye ZA. COVID-19 associated multisystem inflammatory syndrome: A systematic review and meta-analysis. Vol. 19, Iranian Journal of Allergy, Asthma and Immunology. Tehran University of Medical Sciences; 2020. p. 570–88.

18. Ruvinsky S, Voto C, Roel M, Fustiñana A, Veliz N, Brizuela M, et al. Multisystem Inflammatory Syndrome Temporally Related to COVID-19 in Children From Latin America and the Caribbean Region: A Systematic Review With a Meta-Analysis of Data From Regional Surveillance Systems. Vol. 10, Front Pediatr. Frontiers Media S.A.; 2022. p. 871765.

19. Hoste L, Paemel R van, Haerynck F. Multisystem inflammatory syndrome in children related to COVID-19: a systematic review. Eur J Pediatr [Internet]. 2021;180(7):2019–34. Available from: https://doi.org/10.1007/s00431-021-03993-5

20. Radia T, Williams N, Agrawal P, Harman K, Weale J, Cook J, et al. Multi-system inflammatory syndrome in children & adolescents (MIS-C): A systematic review of clinical features and presentation. Vol. 38, Paediatr Respir Rev. W.B. Saunders Ltd; 2021. p. 51–7.

21. Yasuhara J, Watanabe K, Takagi H, Sumitomo N, Kuno T. COVID-19 and multisystem inflammatory syndrome in children: A systematic review and meta-analysis. Pediatr Pulmonol. 2021 May 1;56(5):837–48.

22. Godfred-Cato S, Abrams JY, Balachandran N, Jaggi P, Jones K, Rostad CA, et al. Distinguishing Multisystem Inflammatory Syndrome in Children from COVID-19, Kawasaki Disease and Toxic Shock Syndrome. Pediatric Infectious Disease Journal. 2022 Apr 1;41(4):315–23.

23. Santos MO, Gonçalves LC, Silva PAN, Moreira ALE, Ito CRM, Peixoto FAO, et al. Multisystem inflammatory syndrome (MIS-C): a systematic review and meta-analysis of clinical characteristics, treatment, and outcomes. Vol. 98, Jornal de Pediatria. Elsevier Editora Ltda; 2022. p. 338–49.

24. Porritt RA, Binek A, Paschold L, Rivas MN, McArdle A, Yonker LM, et al. The autoimmune signature of hyperinflammatory multisystem inflammatory syndrome in children. Journal of Clinical Investigation. 2021;131(20).

25. Antúnez-Montes OY, Escamilla MI, Figueroa-Uribe AF, Arteaga-Menchaca E, Lavariega-Saráchaga M, Salcedo-Lozada P, et al. COVID-19 and Multisystem Inflammatory Syndrome in Latin American Children: A Multinational Study. Pediatr Infect Dis J. 2021;40(1):e1–6.

26. Miller A, Zambrano L, Yousaf AR, Abrams JY, Meng L, Wu MJ. Multisystem Infalmmatory Syndrome in Children-United States, February 2020-July 2021. Clin Infect Dis. 2022;75(1):e1165–75.

27. Belay ED, Abrams J, Oster ME, Giovanni J, Pierce T, Meng L, et al. Trends in Geographic and Temporal Distribution of US Children with Multisystem Inflammatory Syndrome during the COVID-19 Pandemic. JAMA Pediatr. 2021 Aug 1;175(8):837–45.

28. Abrams JY, Oster ME, Godfred-Cato SE, Bryant B, Datta SD, Campbell AP, et al. Factors linked to severe outcomes in multisystem inflammatory syndrome in children (MIS-C) in the USA: a retrospective surveillance study. Lancet Child Adolesc Health. 2021 May 1;5(5):323–31.

